# Association of infected probability of COVID-19 with ventilation rates in confined spaces: a Wells-Riley equation based investigation

**DOI:** 10.1101/2020.04.21.20072397

**Authors:** Hui Dai, Bin Zhao

## Abstract

**Background:** A growing number of epidemiological cases are proving the possibility of airborne transmission of coronavirus disease 2019 (COVID-19). Ensuring adequate ventilation rate is essential to reduce the risk of infection in confined spaces.

**Methods:** We obtained the quantum generation rate by a COVID-19 infector with a reproductive number based fitting approach, and then estimated the association between infected probability and ventilation rate with the Wells-Riley equation.

**Results:** The estimated quantum generation rate of COVID-19 is 14-48 /h. To ensure infected probabolity less than 1%, ventilation rate lareger than common values (100-350 m^3^/h and 1200-4000 m^3^/h for 15 minutes and 3 hours exposure, respectively) is required. If both the infector and susceptibles wear masks, the ventilation rate ensuring less than 1% infected probability is reduced to 50-180 m^3^/h and 600-2000 m^3^/h correspondingly, which is easier to be achieved by normal ventilation mode applied in some typical scenarios, including offices, classrooms, buses and aircraft cabins.

**Interpretation:** The risk of potential airborne transmission in confined spaces cannot be ignored. Strict preventive measures that have been widely adopted should be effective in reducing the risk of airborne transmitted infection.

## 1. Introduction

The once-in-a-century coronavirus disease 2019 (COVID-19) pandemic shows that the risk of infection in public confined spaces cannot be ignored. Although the transmission of COVID-19 occurs mainly via droplets during close contact or contaminated surfaces, a recent study showed that severe acute respiratory syndrome coronavirus 2 (SARS-CoV-2) remains viable in aerosols for multiple hours [1], and a latest report issued by the National Academies of Sciences, Engineering, and Medicine suggested that currently available research supports the possibility that SARS-CoV-2 could be spread via bioaerosols generated directly by patients’ exhalation based on collected evidences [2]. A latest field sampling study further indicates that SARS-CoV-2 was widely distributed in the air and the transmission distance in the air might be up to 4 m [3]. There have been many studies suggest that insufficient ventilation increases disease transmission [4]. Therefore, ensuring the safety of the ventilation and air conditioning system for offices, classrooms and public transport is essential to reduce the risk of infection in these confined spaces, which is extremely important for our daily life.

In this study, we employed the Wells–Riley equation [5] to estimate the association between infected probability and ventilation rate. We firstly obtained the quantum generation rate by a COVID-19 infector with a fitted correlation between reproductive number and quantum generation rate, and then estimated association between infected probability and ventilation rate for some typical scenarios, including offices, classrooms, buses and aircraft cabins.

## 2. Method

### 2.1 Wells-Riley Equation

The Wells–Riley equation is as follows [5]:

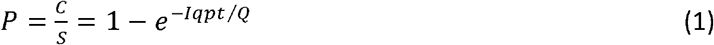

Where *P* is the probability of infection (risk). *C* is the number of cases to develop infection, *S* is the number of susceptibles. *I* is the number of source patients (infector). In this study, we focused on normal scenarios (not health care settings for COVID-19 patients), thus we assumed *I*=1. p is the pulmonary ventilation rate of each susceptible per hour (m^3^/h), *p*=0.3 m^3^/h when people sitting or doing light activity indoor [6]; *Q* is the room ventilation rate (m^3^/h); q is the quantum generation rate produced by one infector (quantum/h); and t is the exposure time (h).

The Wells-Riley equation has the key assumptions: (1) Droplet nuclei are evenly distributed in space, which means the infection risk predicted by this equation is uniform within the space; (2) The equation neglects viability and infectivity of the pathogen quanta.

As for the effect of wearing mask, we consider wearing mask is equivalent to increasing the room ventilation rate due to the filtration effect of the mask, resulting a diluted concentration that susceptibles inhale. The filtration efficiency of ordinary medical surgical mask on virus-laden aerosols is assumed to be 60% [7]. Considering the influence of air leakage, the filtration efficiency can be considered as 50% [8], which is equivalent to double the ventilation rate. If both the infectors and susceptibles wear masks, the ventilation rate is increased to 4 times equivalently.

The key input parameter when applying the Wells–Riley equation is quantum generation rate, *q*, which is determined by the infectious disease we concern.

### 2.2 Quantum generation rate with a reproductive number based fitting approach

So far, there have been no available literatures reported the quantum generation rate (q) of COVID-19. To obtain a reasonable quantum generation rate of COVID-19 for applying the Wells–Riley equation, we collected the known quantum generation rate (q) and basic reproductive number (*R*_*0*_) for other airborne transmitted infectious diseases in previous studies, and fitted the association between *q* and *R*_*0*_. Then we estimated *q* with *R*_*0*_ of the COVID-19 based on the fitted equation. The basic reproductive number is the key epidemiological determinant that characterizes the transmission potential of a certain infectious disease, which is defined as the average number of infectious individuals created by a single infectious person in a completely susceptible population [9]. As basic reproductive number essentially determines the rate of spread of an epidemic, thus we think it may be logical and reasonable to obtain quantum generation rate based on its association with basic reproductive number. Table 1 lists the values of q and R_0_ for several typical infectious diseases collected from references. With the values listed in this table as inputs, we fitted the association between *q* and *R*_*0*_ with a least square method implemented in Origin 2019

**Table 1.** *q* and *R*_*0*_ of airborne transmitted infectious diseases

| | $q(/h)$ | $R_0$ |
| --- | --- | --- |
| **Tuberculosis** | 1-50 [10] | 2.22-5.46 [11] |
| **MERS** | 6-140 [10] | <1(1.0-5.7) [12] |
| **SARS** | 10-300 [10] | 2-5 [13] |
| **Influenza** | 15-500 [10] | 1.6-3.0 [14] |
| **Measles** | 570-5600 [10] | 11-18 [15] |

## 3. Results

### 3.1 The fitted quantum generation rate of COVID-19 with *R*_*0*_

The fitted curve representing the association between *q* and *R*_*0*_ is shown in Figure 1, and the fitted equation is:

**Figure 1.**
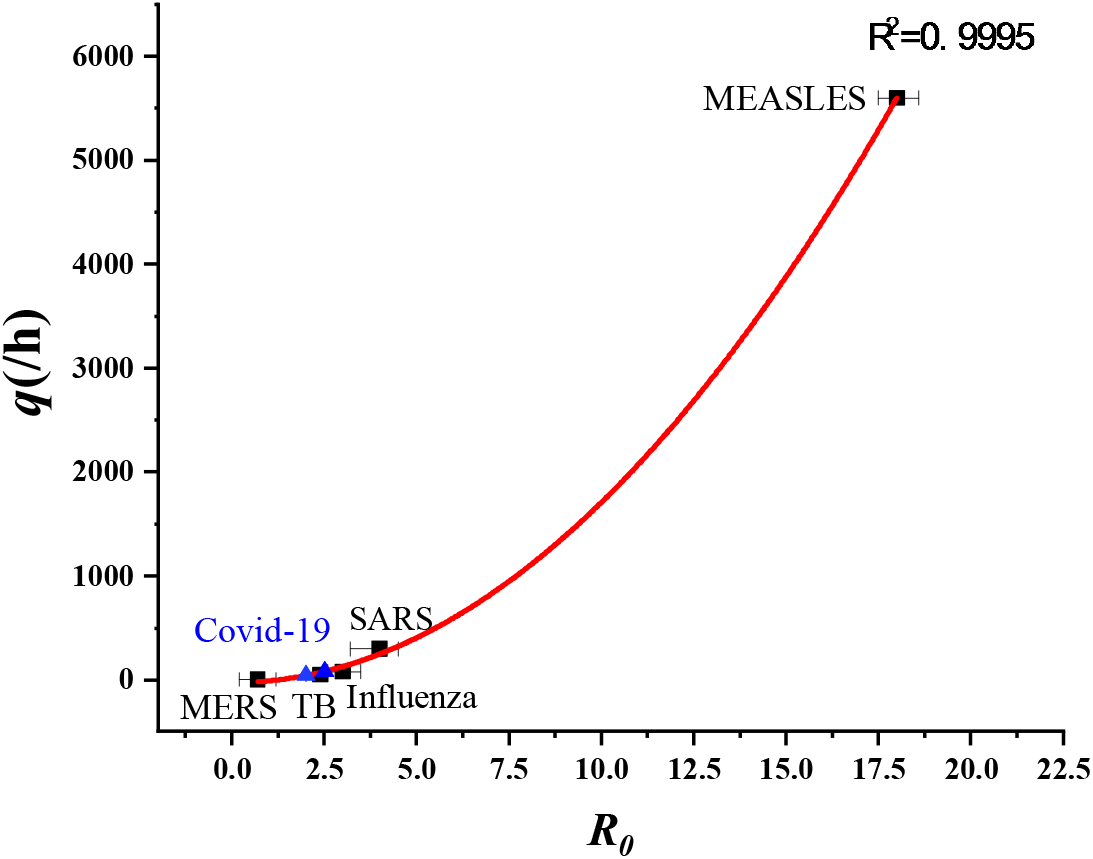
The fitted curve between *q* and *R*_*0*_.

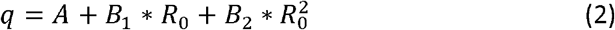

where *A*= −26.06428±68.60109, *B*_1_= −0.61896±29.44301, *B*_2_=17.40102±1.46132. With the widely used range of *R*_0_ of 2-2.5[16-19], we obtained the corresponding range of *q* for COVID-19, which is 14 - 48/h.

### 3.2 Association between infected probability and ventilation rate

#### Infection probability

Figure 2 shows the estimated association between infected probability and ventilation rate. For the case there is one infector inside the confined space, 100-350 m^3^/h of ventilation rate is required to ensure susceptibles exposed for 15 minutes with less than 1% infected probabolity; and 1200-4000 m^3^/h of ventilation rate is required to ensure infected probabolity less than 1% for 3 hours-exposured susceptibles.

**Figure 2.**
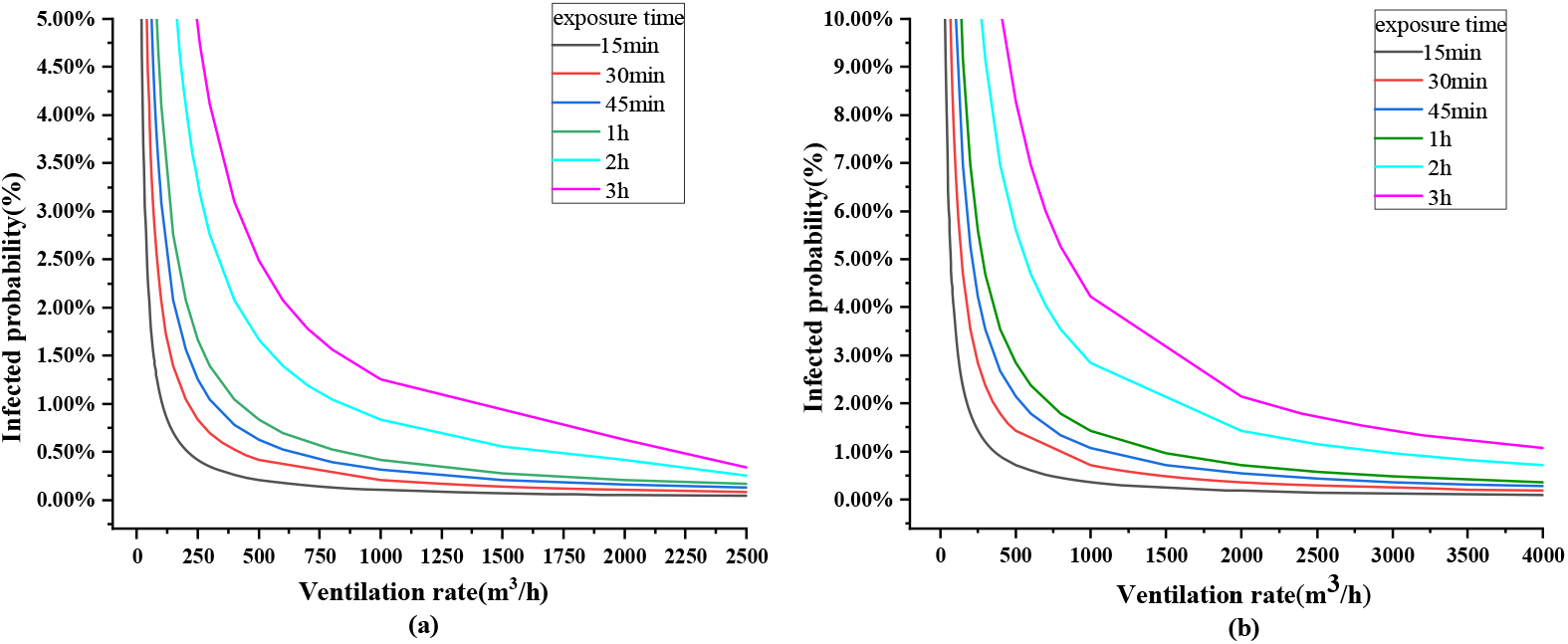
Infected probability with different ventilation rate during different exposure time (no mask) (a) *q*=14 /h (b) *q*=48.

#### The role of wearing mask

The estimated association between infected probability and ventilation rate for the wearing mask cases is shown in Figure 3. It indicates wearing mask play an important role in reducing the probability of infection. For the scenario where there is one infector in the space, if both the infector and susceptibles wear masks, the required ventilation rate to ensure infected probability less than 1% is reduced to 50-180 m^3^/h for 15 min-exposured susceptibles, and 600-2000 m^3^/h for 3h-exposured susceptibles, respectively.

**Figure 3.**
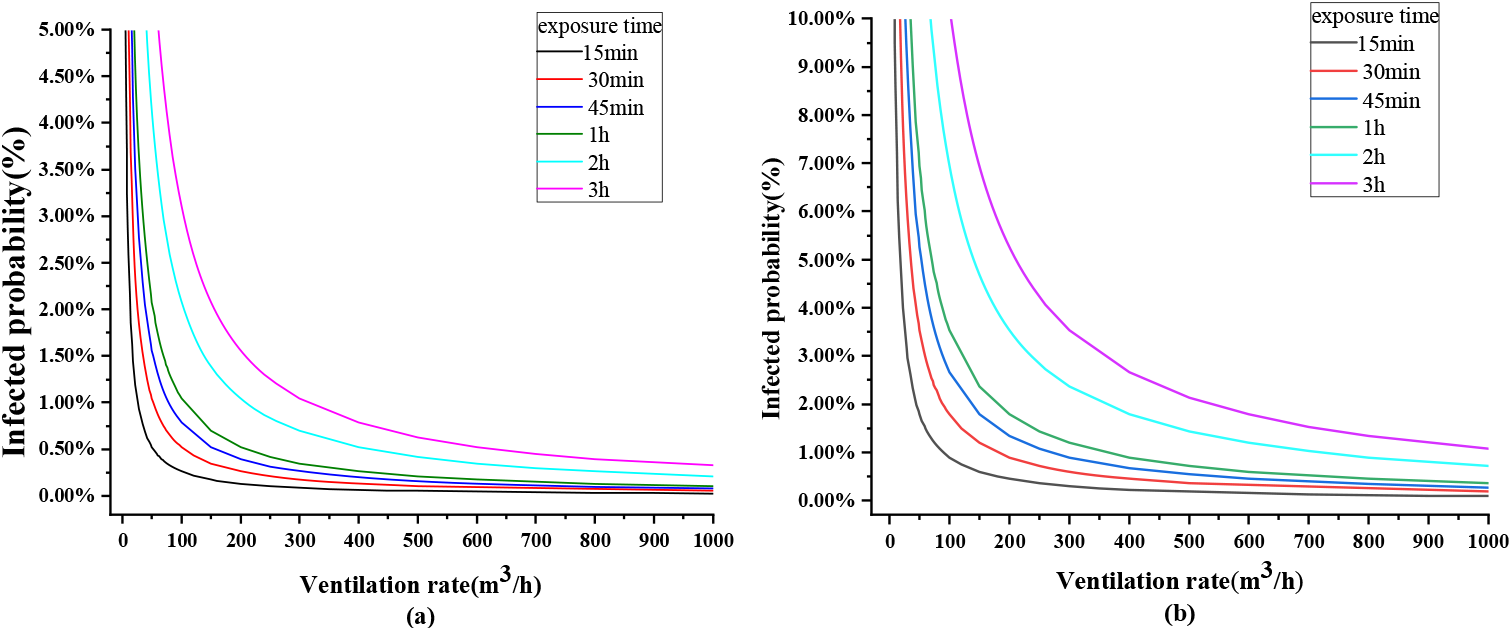
Infected probability with different ventilation rate during different exposure time (a) *q*=14 /h (b) *q*=48 /h (with mask)

#### Some typical scenarios

Figure 4 shows the estimated infected probability association with ventilation rate for some typical scenarios. Table 2 lists the corresponding air change rates (ACH). The air change rate is a measurement of how much fresh/clean air replaces indoor air in one hour [20], which is easy to applied to judge if the actual ventilation and air conditioning system could provide enough ventilation rate to ensure low infected probability.

**Table 2.** ACH vs. infected probability (Y: with mask; N: no mask)

| Infected probability | Scenarios | Bus |  | Classroom |  | Aircraft cabin |  | Office |  |
| --- | --- | --- | --- | --- | --- | --- | --- | --- | --- |
|  | Volume (m<sup>3</sup>) | 75 |  | 348 |  | 100 |  | 150 |  |
|  | Exposure time (h) | 0.5 |  | 2 |  | 4 |  | 8 |  |
|  |  | ACH (/h) |  |  |  |  |  |  |  |
|  |  | Y | N | Y | N | Y | N | Y | N |
| 2.0% | *q*=14 /h | 0.33 | 1.3 | 0.28 | 1.15 | 2 | 8 | 2.7 | 10 |
| 1.5% |  | 0.48 | 1.8 | 0.4 | 1.7 | 3 | 11 | 3.3 | 14 |
| 1.0% |  | 0.7 | 2.8 | 0.6 | 2.4 | 4 | 16 | 6 | 20 |
| 0.5% |  | 1.4 | 5.6 | 1.2 | 4.8 | 9 | 33 | 10 | 43 |
| 0.1% |  | 7 | 27 | 5.5 | 20 | 35 | 160 | 50 | 200 |
| 2.0% | *q*=48 /h | 1.2 | 4.8 | 1 | 4 | 7 | 30 | 10 | 36 |
| 1.5% |  | 1.6 | 6.4 | 1.4 | 5 | 9 | 38 | 12 | 53 |
| 1.0% |  | 2.4 | 9.6 | 2 | 7 | 15 | 55 | 18 | 80 |
| 0.5% |  | 4.8 | 19 | 3.5 | 15 | 29 | 110 | 33 | 133 |
| 0.1% |  | 24 | 93 | 20 | 71 | 125 | 550 | 200 | 666 |

The ACHs in red text indicate the ventilation rates for the corresponding scenarios could be achieved with its normal ventilation system or natural ventilation.

**Figure 4.**
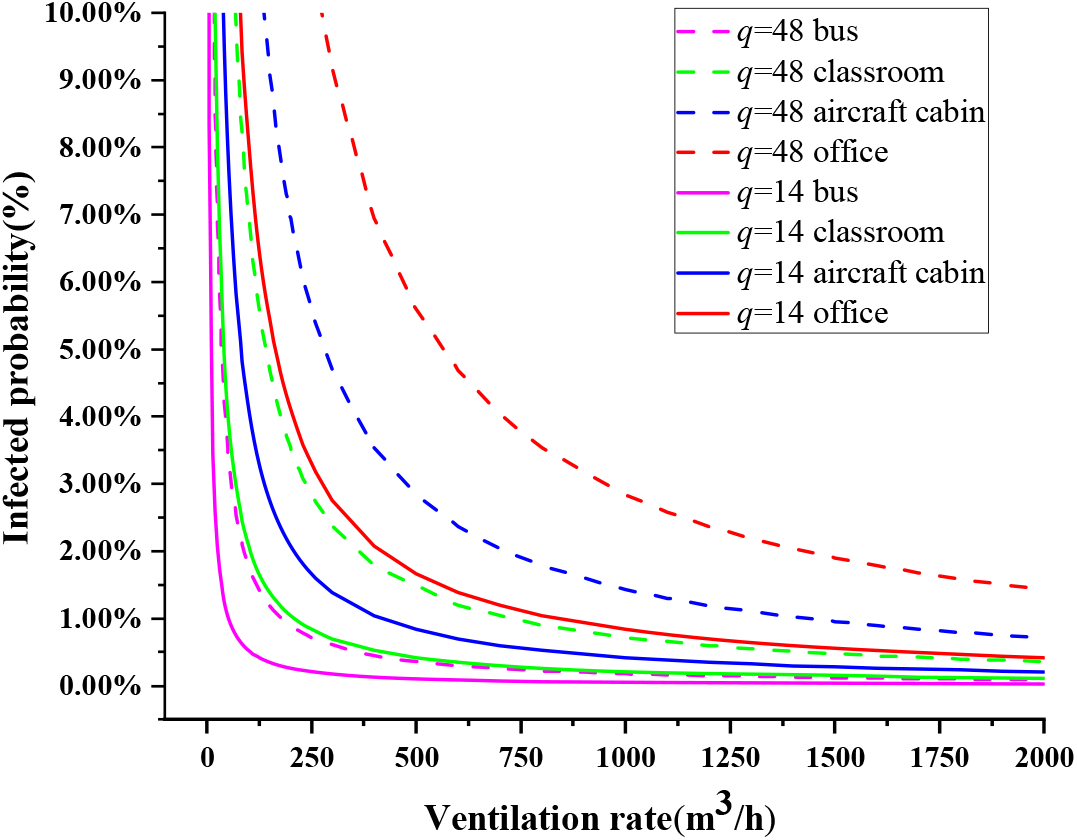
Association between infected probability and ventilation rate in typical.

If people wearing mask, natural ventilation or normal mechanical ventilation can provide enough ventilation rate ensureing infected probability less than 1% for most scenarios (except for office, as shown table 2). However, without wearing mask may result a relative higher infection risk for most cases, especially if *q* reachs the maximum.

## Discussion

The estimation indicates that, once an infector (especially an asymptomatic infector has not been identified as confirmed COVID-19 patient) enters a public confined space, the risk of infection is relative high, with approximately 2% of infected probability at the common ventilation rate (500-2500 m^3^ /h as shown in Figure 2). Such findings may partly explain the early large-scal epidemics in China and the other European and American countries.

Wearing a ordinary medical surgical mask is effective, thus it is important to educate people wearing mask when they enter or stay in confined spaces. Furthermore, the above finding imply that preventing the infectors (especially asymptomatic infectors has not been confirmed as patient) from entering those public spaces is critical to suppressing the spread of the virus via airborne transmission. Current measures, strict screening of asymptomatic infectors with more widely nucleic acid test and home isolation have already ensured that infectors are unlikely to enter public spaces, also ensured a relatively safe environment of confined spaces.

We acknowledge that direct evidences of airborne transmission of COVID-19 are still lack. However, the risk of potential or opportunistic airborne transmission cannot be ignored, and the value of q warrants further study. Also, there is no need to panic. As govements and public have realized the emergency of the pandemic, strict preventive measures have been widely adopted, which are effective in reducing the risk of airborne transmitted infection.

## Data Availability

All data could be provided by the authors if requested.

